# Factors Associated with Overweight and Obesity among Women of Reproductive Age in Cambodia: Analysis of Cambodia Demographic and Health Survey

**DOI:** 10.1101/2023.04.28.23289243

**Authors:** Samnang Um, Yom An

## Abstract

Overweight and obesity are associated with increased chronic disease and death rates globally. In Cambodia, the prevalence of overweight and obesity among women is high and may grow. This study aimed to determine the prevalence and factors associated with overweight and obesity among women of reproductive age (WRA) in Cambodia. We analyzed data from the 2021-2022 Cambodia Demographic and Health Survey (CDHS) that used a two-stage stratified cluster sampling design. Data analysis was restricted to non-pregnant women, resulting in an analytic sample of 9,417 WRA. Multivariable logistic regressions were performed using STATA V17 to examine factors associated with overweight and obesity. Prevalence of overweight and obesity among non-pregnant women of reproductive age was 22.56%, and 5.61% were overweight and obese, respectively. Factors independently associated with increased odds of overweight and obesity included women aged 20-29 years [AOR=1.85; 95% CI: 1.22 - 2.80], 30-39 years [AOR=3.34; 95% CI: 2.21 - 5.04], and 40-49 years [AOR=5.57; 95% CI: 3.76 - 8.25], married women [AOR=2.49; 95% CI: 1.71-3.62], women from middle wealth quintile [AOR=1.21; 95% CI: 1.02-1.44], and rich wealth quintile [AOR=1.44; 95% C: 1.19 - 1.73], having at least three children or more [AOR=1.40; 95% CI: 1.00 - 1.95], ever drink alcohol [AOR=1.24; 95% CI: 1.04 - 1.47], and current drink alcohol [AOR=1.2; 95% CI: 1.01 - 1.45]. On the contrary, the following factors were independently associated with decreased odds of having overweight and obese: women with at least secondary education [AOR=0.73; 95% CI: 0.58-0.91] and working in manual labor jobs [AOR=0.76; 95% CI: (0.64 - 0.90]. Increased age, married women, having at least three children, and alcohol consumption were the main risk factors associated with overweight and obesity. Conversely, higher education and manual labor were negatively associated with overweight and obesity. Cambodia’s non-communicable disease (NCD) public health programs should consider these characteristics for targeting interventions further to reduce overweight and obesity in the coming years.

## Introduction

Overweight and obesity are significant global public health challenges that have rapidly risen in the last four decades and are regarded as an epidemic [1]. In 2016, approximately 39% (1.9 billion) of adults aged 18 years and older were overweight, and 13% (650 million) were obese worldwide [1]. Being overweight and obese are major risk factors for several non-communicable diseases (NCDs), including cardiovascular (CVDs) and kidney diseases, type 2 diabetes, some cancers, musculoskeletal disorders, and other chronic diseases [2,3]. Moreover, among women of reproductive age (WRA), overweight and obesity have been associated with increased risk of pregnancy complications, cesarean section births, adverse birth outcomes, and infant mortality [4]. Overweight and obesity are the fourth leading cause of risk-attributable mortality [5], with a reduced life expectancy of 5-20 years, depending on the condition’s severity and comorbidities [1]. According to the WHO, being overweight and obese are the leading risks for global deaths, with at least 2.8 million adults dying annually due to these conditions [1]. Overweight and obese were higher among women than men in both developed and developing countries; for instance, overweight was 40% in women vs. 39% in men and obesity 15% in women vs. 11% in men [1]. The Global Nutrition Report 2019 showed that 26.1% of women and 20.4% of men were overweight, while obesity was 6.3% in women compared to 3.5% in men [6]. Along with the rapidly increasing population in Cambodia, from 15.42 million in 2015 to nearly 16.21 million in 2019 [7], the prevalence of overweight and obesity is also steadily rising. According to the Cambodia STEPS survey indicate the overall prevalence of currents smoking any tobacco was 29.4% in 2010 to 22.1% in 2016, and currents drinking alcohol was 53.5% in 2010 to 46% in 2016, 11% were low physical activity in 2010 to 13% in 2016 [11]. In addition, 63% of women consumed sweet beverages, and 33% consumed unhealthy foods the previous day in 2021-22 [8]. The 2021-22 Cambodia Demographic and Health Survey (CDHS) report indicated that overweight and obesity among non-pregnant women of reproductive age increased from 18% (15.2% overweight and 2.8% obese) in 2014 to 33% (26% overweight and 6% obese) in 2021-22 [8]. It was estimated that overweight and obese contribute to rising healthcare costs in Cambodia (approximately 1.7% of its annual gross domestic product (GDP) per capita) and is a significant contributor to mortality and decreased general health and productivity [9]. Predictors of overweight and obesity among WRA from Demographic and Health Survey (DHS) data such as Cambodia, Bangladesh, Nepal, India, and Ethiopia include higher socioeconomic status, older age, marriage, living in an urban residence, and lack of education [3,10-12]. Women with formal employment had higher odds of being overweight or obese than informally employed women [3,13]. Also, women who used hormonal contraceptives such as oral contraceptive pills, implants, patches, and rings were at higher risk of being overweight or obese [14,15]. Globally, 30% of daily smokers are overweight or obese [16], with women smokers at greater risk for obesity than men smokers [17,18]. Women with frequent television watching [19], alcohol drinking [20,21], and frequent consumption of sweets foods and unhealthy foods [22] were found to have higher odds of being overweight or obese. To our knowledge, factors associated with overweight and obesity, specifically among WRA in Cambodia using updated data, have not been explored. A previous study on the prevalence of overweight and obesity among WRA and its associated factors utilized data since 2014 [3]. In Cambodia, where the prevalence of overweight and obesity in WRA has increased rapidly, a comprehensive investigation of a wide range of socio-demographic and behavioral factors is warranted to identify the factors independently associated with having overweight and obesity. Identifying the critical modifiable socio-demographic and behavioral factors, as well as women at a high risk of having overweight and obese, may help guide the timely development of promising and feasible public health intervention strategies to address the growing overweight and obesity pandemic. Therefore, we aimed to determine the prevalence and examined socio-demographic and behavioral factors associated with overweight and obesity among WRA in Cambodia.

## Material and Methods

### Ethics statement

The data used in this study were extracted from CDHS 2021-22 data [8], which are publicly available with all personal identifiers of study participants removed. Also, the CDHS data are publicly accessible and were made available to us upon request through the DHS website at (URL: https://dhsprogram.com/data/available-datasets.cfm). Written informed consent was obtained from the parent/guardian of each participant under 18 years of age. Also, the study contains no individual identifiers that could affect the confidentiality of the participants, and the data were used for analysis purposes only. The data collection tools and procedures for CHDS 2021-22 were approved by the Cambodia National Ethics Committee for Health Research 10 May 2021 (**Ref: 083 NECHR**) and the Institutional Review Board (IRB) of ICF in Rockville, Maryland, USA.

### Data sources and procedures

We followed the methods of **Um S et al**., **2023** [3]. To analyze the prevalence and factors associated with overweight and obesity among WRA in Cambodia, we used existing women’s data from the 2021-22 CDHS. The CDHS is a nationally representative population-based household survey implemented by the National Institute of Statistics (NIS) in collaboration with the Ministry of Health (MoH) with financial support from the Government of Cambodia and technical assistance from ICF International (USA) through the Demographic and Health Survey program. Data collection was conducted from September 15, 2021, to February 15, 2022 [8]. The sampling frame used for the 2021-22 CDHS was taken from the Cambodia General Population Census 2019 [7]. After that, the CDHS 2021-2022 participants were selected using probability proportion based on two-stage stratified cluster sampling from the chosen sampling frame. In the initial stage, 709 enumeration areas (EAs) (241 urban areas and 468 rural areas) were selected. In the second stage, an equal systematic sample of 30 households was selected from each cluster for a total sample size of 21,270 households. In total, 19,496 women aged 15-49 were interviewed, with a response rate of 98.2%. Survey data were obtained through face-to-face interviews using a standardized survey instrument by trained interviewers. Anthropometric weight and height measurements for adult women aged 15-49 were taken by trained female field staff using standardized instruments and procedures [7]. Weight measurements were taken using scales with a digital display (UNICEF model S0141025). Height and length were measured using a portable adult height measurement system (UNICEF model S0114540). The detailed protocol and methods were published previously [8]. Eligible participants for this study were women of reproductive age 15-49 years, with the exclusion of pregnant women and those who had a birth within two months before the survey, with available body mass index (BMI) data. As a result, we excluded 828 pregnant women and 9,249 women with missing BMI data. A final sample included in this analysis was 9,417 women of reproductive age 15-49 years.

## Measurements

### Outcome variable

The primary outcome variable of this study was overweight and obese for women of reproductive age. BMI - defined by dividing a person’s weight in kilograms by the square of their height in meters (kg/m^2^) – was used to measure the outcome [8,23]. For adults over 15 years old, the following BMI ranges were used: Underweight (<□18.0□kg/m^2^), Normal weight (18.5-24.9 kg/m^2^), Overweight (25.0-29.9 kg/m^2^), and Obesity (≥ 30.0 kg/m^2^) [8,23]. **Overweight/Obese** was defined as a binary outcome for which women with a BMI ≥ 25.0 kg/m^2^ were classified as overweight and obese (coded = 1), while women with a BMI < 25.0 kg/m^2^ were coded as other (coded = 0).

### Covariate variables

Covariates included sociodemographic characteristics of women of reproductive age: Women’s age in years (15-19, 20-29, 30-39, and 40-49), marital status (not married, married or living together, and divorced or widowed or separated), educational level (no formal education, primary, secondary or higher education), occupation (not working, agriculture, manual labor or unskilled, professional or sealer or services), number of children ever born (no children, one-two child, and three and more children). Households’ wealth status was represented by a wealth index calculated via principal component analysis (PCA) and using variables for household assets and dwelling characteristics. Weighted scores are divided into five wealth quintiles (poorest, poorer, medium, richer, and richest), each comprising 20% of the population [8] and place of residence (rural vs. urban). Cambodia’s provinces were regrouped for analytic purposes into a categorical variable with four geographical regions: plains, Tonle Sap, coastal/sea, and mountains [7]. Behavioral factors including smoking (non-smoker vs. smoker), alcohol consumption in the past month (never drink, ever drink, and current drink), current drink of alcohol correspond to one can or bottle of beer, one glass of wine, or one shot of spirits in the past month, watching television at least once a week (yes vs. no), contraceptive usage (not used, hormonal methods (using a pill, emergency contraceptive pill, Norplant, injection, and vaginal rings), non-hormonal methods (condoms, the diaphragm, the IUD, spermicides, lactational amenorrhea, and sterilization), and traditional methods (periodic abstinence and withdrawal) [8].

### Statistical analysis

All statistical analyses were performed using STATA version 17 (Stata Corp 2021, College Station, TX) [24], accounting for the CDHS sampling weights and complex survey design. Socio-demographic characteristics and behavioral factors were described in weighted frequency and percentage. The provincial variation in the prevalence of overweight and obesity was done using ArcGIS software version 10.8 [24]. Shapefiles for administrative boundaries in Cambodia are publicly accessible through the DHS website (URL: https://spatialdata.dhsprogram.com/boundaries/#view=table&countryId=KH).

Descriptive analyses that coupled cross-tabular frequency distributions with bivariate chi-square tests were used to assess associations between explanatory variables and the outcome variable Overweight/Obese. Variables associated with the outcome variable with a significance level of p-value ≤ 0.10 [3] and follow these presents study [25,26] as potential confounder variables (for example, women’s age, wealth index, alcohol consumption, residence, and geographic region) were included in the adjusted logistic regression analyses. Simple logistic regression was used to determine the magnitude effect of associations between overweight and obesity with socio-demographic characteristics and behavioral factors reported as odds ratios (OR) with 95% confidence intervals (CI). Then, Multiple logistics regression was used to assess independent associations, reported as adjusted odds ratios (AOR), with overweight and obesity after adjusting for other independent variables included in the model. Multicollinearity between independent variables was checked before fitting the final regression model (see S1 Table).

## Results

### Characteristics of the study samples

The mean age of women was 31 years old (SD = 9.6 years), in which the age group 30-49 years old accounted for 57.11%. Almost 67.82% were married, 49.47% had completed at least secondary education, and 11.90% did not receive formal schooling. 30.30% of women did professional work, and 25.86% were unemployed. Of the total sample, 35.97% of women were from poor households. Over half (57.27%) of women resided in rural areas. About 27.79% had three and more children, while 29.49% had no children. Only 1.40% of women reported cigarette smoking, 16.60% reported currently drinking alcohol, 18.6% reported ever drinking, and 12.35% reported using hormonal contraceptives. The mean women’s BMI was 22.9 kg/m^2^ (SD = 3.9 kg/m^2^), and 22.56% and 5.61% were overweight and obese, respectively (see **Table 1**).

**Table 1.**
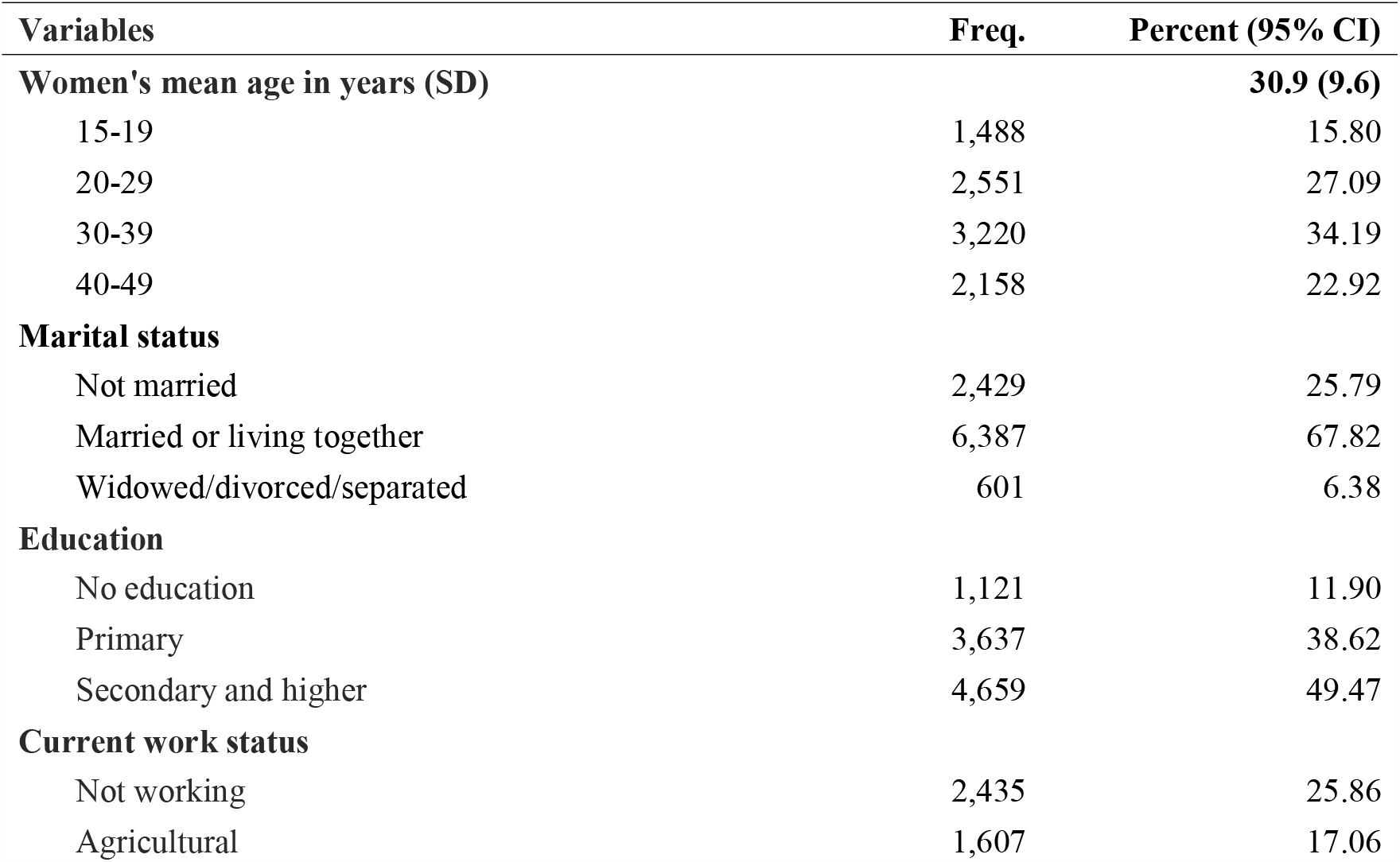
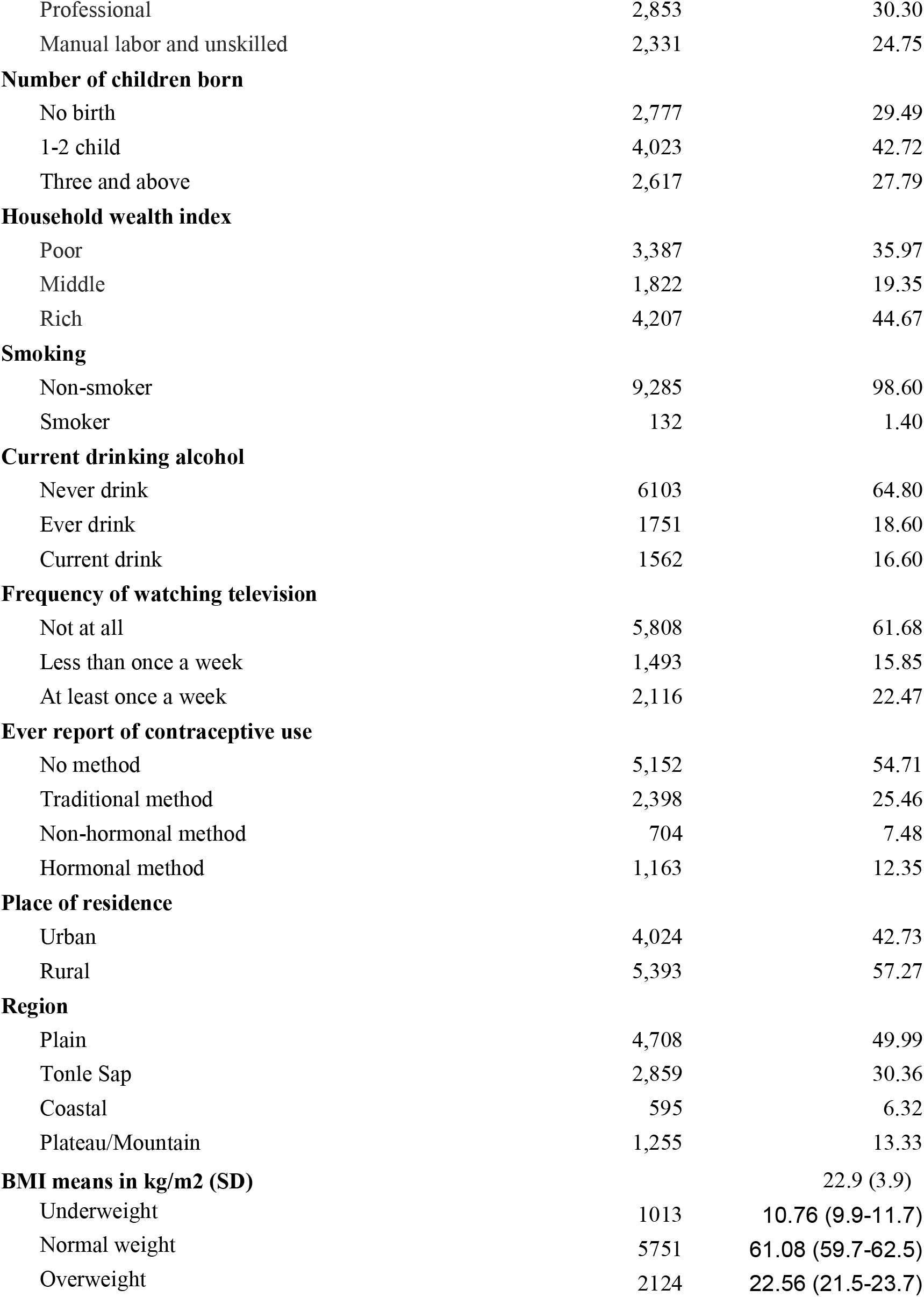
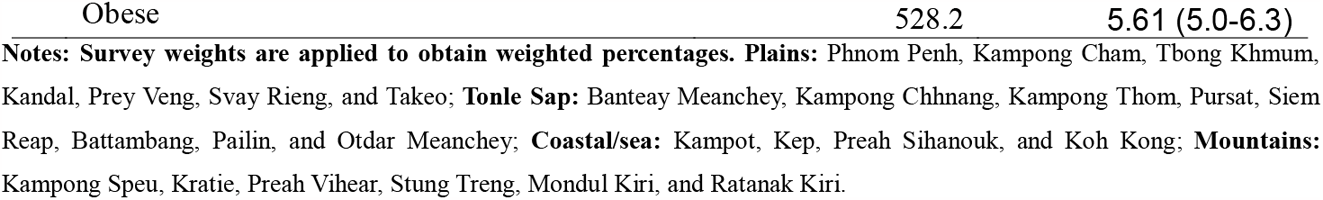
Socio-demographic and behaviors characteristics of the weighted samples of women aged 15-49 years old in Cambodia, 2021-2022 (n□ =□9,417, weighted count)

### Prevalence of overweight and obesity among WRA by provinces

The prevalence of overweight and obesity is highest among WRA in Kampong Cham (34.1%), Kandal (32.3%), Svay Rieng (32%), Preah Sihanouk (31.2%), Phnom Penh, Kompong Thom, Pailin, and Tboung Khmum (30% each) and lowest among WRA in Ratanak Kiri (9.4%) and Kampong Chhnang (14.6%) (**see Fig 1, and S2 Table**).

**Fig 1.**
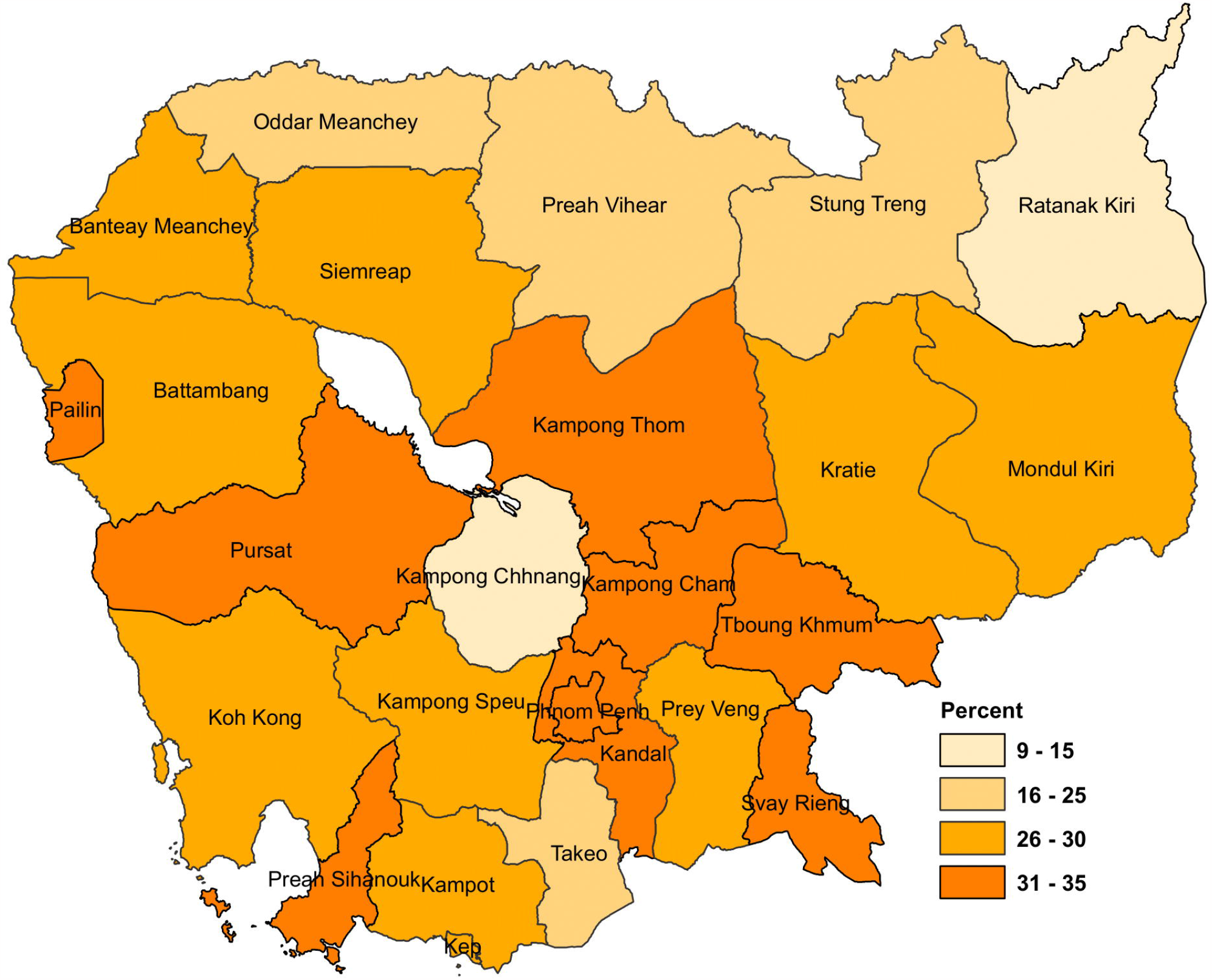
Prevalence of overweight and obesity in Women Reproductive Age by province. Map created using ArcGIS software version 10.8 [24]. Shapefiles for administrative boundaries in Cambodia are publicly accessible through the DHS website (URL: https://spatialdata.dhsprogram.com/boundaries/#view=table&countryId=KH).

### Factors associated with overweight and obesity in bivariate analysis

Socio-demographic characteristics and behavioral factors were significantly associated with overweight and obesity (**Table 2**). Women had a higher prevalence of overweight/obesity if they were aged 40-49 years (47.31%) compared to younger age groups (p-value <0.001), married (35.87%) compared to another relationship status (p-value <0.001), had no education (37.91%) compared to higher education (p-value <0.001), were employed in a professional job (34.03%) compared to other jobs, or were from the rich households (30.90%) compared to poor households (p-value <0.001). In addition, Overweight/obesity increased with parity. Women reported at least three children had a significantly highest overweight and obesity (42.53%) (p-value < 0.001). Women who currently drink alcohol had a significantly higher proportion of overweight and obese (33.87%) than women who did not (p-value <0.001). Women who used hormonal contraceptive methods had a significantly higher proportion of overweight and obesity (37.92%) than women not use contraceptive (p-value <0.001). Geographic regions of residence were likewise associated with a woman having overweight and obese. Women who lived in urban areas had a higher prevalence of overweight and obesity than those living in rural areas (29.90% vs 26.87%, p-value = 0.028). Plain regions were positively associated with overweight and obesity (29.91%) compared to other regions (p-value = 0.006).

**Table 2.** Factors associated with overweight and obesity among women aged 15-49 years in Chi^2^ analysis (n□=□9,417, weighted count)

| Characteristics | Overweight/Obese<br>(n=2,652) |  | Normal/Underweight<br>(n= 6,764) |  | P value |
| --- | --- | --- | --- | --- | --- |
|  | Freq. | % | Freq. | % |  |
| **Women's age group in years** |  |  |  |  |  |
| 15-19 | 80 | 5.38 | 1,408 | 94.62 | **<0.001** |
| 20-29 | 444 | 17.40 | 2,107 | 82.60 |  |
| 30-39 | 1,108 | 34.41 | 2,112 | 65.59 |  |
| 40-49 | 1,021 | 47.31 | 1,137 | 52.69 |  |
| **Marital status** |  |  |  |  |  |
| Not married | 181 | 7.45 | 2,248 | 92.55 | **<0.001** |
| Married or living together | 2,291 | 35.87 | 4,096 | 64.13 |  |
| Widowed/divorced/separated | 181 | 30.12 | 420 | 69.88 |  |
| **Education** |  |  |  |  |  |
| No education | 425 | 37.91 | 695 | 62.00 | **<0.001** |
| Primary | 1,254 | 34.48 | 2,383 | 65.52 |  |
| Secondary and higher | 973 | 20.88 | 3,686 | 79.12 |  |
| **Current work status** |  |  |  |  |  |
| Not working | 586 | 24.07 | 1,849 | 75.93 | **<0.001** |
| Agricultural | 485 | 30.18 | 1,121 | 69.76 |  |
| Professional | 971 | 34.03 | 1,881 | 65.93 |  |
| Manual labor and unskilled | 563 | 24.15 | 1,768 | 75.85 |  |
| **Number of children born** |  |  |  |  |  |
| No birth | 264 | 9.51 | 2,512 | 90.46 | **<0.001** |
| 1-2 child | 1,275 | 31.69 | 2,748 | 68.31 |  |
| Three and above | 1,113 | 42.53 | 1,504 | 57.47 |  |
| **Household wealth index** |  |  |  |  |  |
| Poor | 841 | 24.83 | 2,546 | 75.17 | **<0.001** |
| Middle | 512 | 28.10 | 1,311 | 71.95 |  |
| Rich | 1,300 | 30.90 | 2,907 | 69.10 |  |
| **Smoking** |  |  |  |  |  |
| Non-smoker | 2,621 | 28.23 | 6,664 | 71.77 | 0.325 |
| Smoker | 32 | 24.24 | 100 | 75.76 |  |
| **Current drinking alcohol** |  |  |  |  |  |
| Never drink | 1,573 | 25.77 | 4,530 | 74.23 | **<0.001** |
| Ever drink | 550 | 31.41 | 1,201 | 68.59 |  |
| Current drink | 529 | 33.87 | 1,033 | 66.13 |  |
| **Frequency of watching television** |  |  |  |  |  |
| Not at all | 1,623 | 27.94 | 4,186 | 72.07 | 0.901 |
| Less than once a week | 423 | 28.33 | 1,069 | 71.60 |  |
| At least once a week | 606 | 28.64 | 1,509 | 71.31 |  |
| **Ever report of contraceptive use** |  |  |  |  |  |
| No method | 1,132 | 21.97 | 4,020 | 78.03 | **<0.001** |
| Traditional method | 821 | 34.24 | 1,577 | 65.76 |  |
| Non-hormonal method | 259 | 36.79 | 446 | 63.35 |  |
| Hormonal method | 441 | 37.92 | 722 | 62.08 |  |
| **Place of residence** |  |  |  |  |  |
| urban | 1,203 | 29.90 | 2,821 | 70.10 | **0.028** |
| rural | 1,449 | 26.87 | 3,944 | 73.13 |  |
| **Region** |  |  |  |  |  |
| Plain | 1,408 | 29.91 | 3,299 | 70.07 | **0.006** |
| Tonle Sap | 768 | 26.86 | 2,092 | 73.17 |  |
| Coastal | 171 | 28.74 | 425 | 71.43 |  |
| Plateau/Mountain | 306 | 24.38 | 949 | 75.62 |  |

**Notes:** Survey weights are applied to obtain weighted percentages. **Plains:** Phnom Penh, Kampong Cham, Tbong Khmum, Kandal, Prey Veng, Svay Rieng, and Takeo; **Tonle Sap:** Banteay Meanchey, Kampong Chhnang, Kampong Thom, Pursat, Siem Reap, Battambang, Pailin, and Otdar Meanchey; **Coastal/sea:** Kampot, Kep, Preah Sihanouk, and Koh Kong; **Mountains:** Kampong Speu, Kratie, Preah Vihear, Stung Treng, Mondul Kiri, and Ratanak Kiri.

### Factors associated with overweight and obesity in adjusted logistic regression

As shown in **Table 3**, several factors were independently associated with increased odds of having overweight and obesity among women. These factors included age group 20-29 years [AOR=1.85; 95% CI: 1.22 - 2.80], 30-39 years [AOR=3.34; 95% CI: 2.21 - 5.04], and 40-49 years [AOR=5.57; 95% CI: 3.76 - 8.25] married [AOR=2.49; 95% CI: 1.71 - 3.62] and widowed/divorced/separated [AOR=1.14; 95% CI: 1.14 - 2.63], middle wealth quintile [AOR=1.21; 95% CI: 1.02 - 1.44], and rich wealth quintile [AOR=1.44; 95% CI: 1.19 - 1.73], having at least three children or more [AOR=1.40; 95% CI: 1.00 - 1.95], ever drink alcohol [AOR=1.24; 95% CI: 1.04 - 1.47], and current drink alcohol [AOR=1.21; 95% CI: 1.01 - 1.45]. On the contrary, the following factors were independently associated with decreased odds of having overweight and obese: women with at least secondary education [AOR=0.73; 95% CI: 0.58-0.91], working in manual labor jobs [AOR=0.76; 95% CI: (0.64 - 0.90] (**see Fig 2)**.

**Table 3.** Risk factors associated with overweight and obesity in unadjusted and adjusted logistic regression analysis.

| Characteristics | Unadjusted (n=9,417) |  | Adjusted (n= 9,225) |  |
| --- | --- | --- | --- | --- |
|  | OR | 95%CI | AOR | 95%CI |
| **Women's age group in years** |  |  |  |  |
| 15-19 | Ref. |  | Ref. |  |
| 20-29 | 3.71*** | (2.61 - 5.27) | **1.85\*\*\*** | **(1.22 - 2.80)** |
| 30-39 | 9.25*** | (6.70 - 12.77) | **3.34\*\*\*** | **(2.21 - 5.04)** |
| 40-49 | 15.84*** | (11.68 - 21.47) | **5.57\*\*\*** | **(3.76 - 8.25)** |
| **Marital status** |  |  |  |  |
| Not married | Ref. |  | Ref. |  |
| Married or living together | 6.96*** | (5.67 - 8.54) | **2.49\*\*\*** | **(1.71 - 3.62)** |
| Widowed/divorced/separated | 5.34*** | (3.81 - 7.50) | **1.73\*\*** | **(1.14 - 2.63)** |
| **Education** |  |  |  |  |
| No education | Ref. |  | Ref. |  |
| Primary | 0.86 | (0.72 - 1.03) | 0.97 | (0.80 - 1.17) |
| Secondary and higher | 0.43*** | (0.35 - 0.53) | **0.73\*\*\*** | **(0.58 - 0.91)** |
| **Current work status** |  |  |  |  |
| Not working | Ref. |  | Ref. |  |
| Agricultural | 1.37*** | (1.14 - 1.64) | 0.85 | (0.69 - 1.03) |
| Professional | 1.63*** | (1.40 - 1.89) | 1.10 | (0.93 - 1.31) |
| Manual labor and unskilled | 1.00 | (0.85 - 1.19) | **0.76\*\*\*** | **(0.64 - 0.90)** |
| **Number of children born** |  |  |  |  |
| No birth | Ref. |  | Ref. |  |
| 1-2 child | 4.41*** | (3.61 - 5.38) | 1.23 | (0.89 - 1.68) |
| Three and above | 7.03*** | (5.79 - 8.55) | **1.40\*\*** | **(1.00 - 1.95)** |
| **Household wealth index** |  |  |  |  |
| Poor | Ref. |  | Ref. |  |
| Middle | 1.18** | (1.02 - 1.37) | **1.21\*\*** | **(1.02 - 1.44)** |
| Rich | 1.35*** | (1.18 - 1.56) | **1.44\*\*\*** | **(1.19 - 1.73)** |
| **Smoking** |  |  |  |  |
| Non-smoker | Ref. |  |  | - |
| Smoker | 0.81 | (0.52 - 1.24) | - | - |
| **Current drinking alcohol** |  |  |  |  |
| Never drink | Ref. |  | Ref. |  |
| Ever drink | 1.32*** | (1.13 - 1.54) | **1.24\*\*** | **(1.04 - 1.47)** |
| Current drink | 1.47*** | (1.23 - 1.76) | **1.21\*\*** | **(1.01 - 1.45)** |
| **Watching television** |  |  |  |  |
| Not at all | Ref. |  |  |  |
| Less than once a week | 1.02 | (0.85 - 1.23) | - | - |
| At least once a week | 1.04 | (0.89 - 1.21) | - | - |
| **Ever report of contraceptive use** |  |  |  |  |
| No method | Ref. |  | Ref. |  |
| Traditional method | 1.85*** | (1.61 - 2.12) | 1.02 | (0.86 - 1.21) |
| Non-hormonal method | 2.06*** | (1.64 - 2.60) | 0.84 | (0.65 - 1.10) |
| Hormonal method | 2.17*** | (1.75 - 2.69) | 1.01 | (0.81 - 1.26) |
| **Place of residence** |  |  |  |  |
| Rural | Ref. |  | Ref. |  |
| Urban | 1.16** | (1.02 - 1.33) | 1.11 | (0.94 - 1.31) |
| **Region** |  |  |  |  |
| Plain | Ref. |  | Ref. |  |
| Tonle Sap | 1.33*** | (1.11 - 1.58) | 1.17 | (0.95 - 1.43) |
| Coastal | 1.14 | (0.95 - 1.36) | 1.11 | (0.91 - 1.35) |
| Plateau/Mountain | 1.25** | (1.00 - 1.55) | 1.10 | (0.86 - 1.41) |

Notes: Survey weights are applied to obtain weighted percentages. \*\*\* p<0.01, \*\* p<0.05, \* p<0.10.

**Fig 2.**
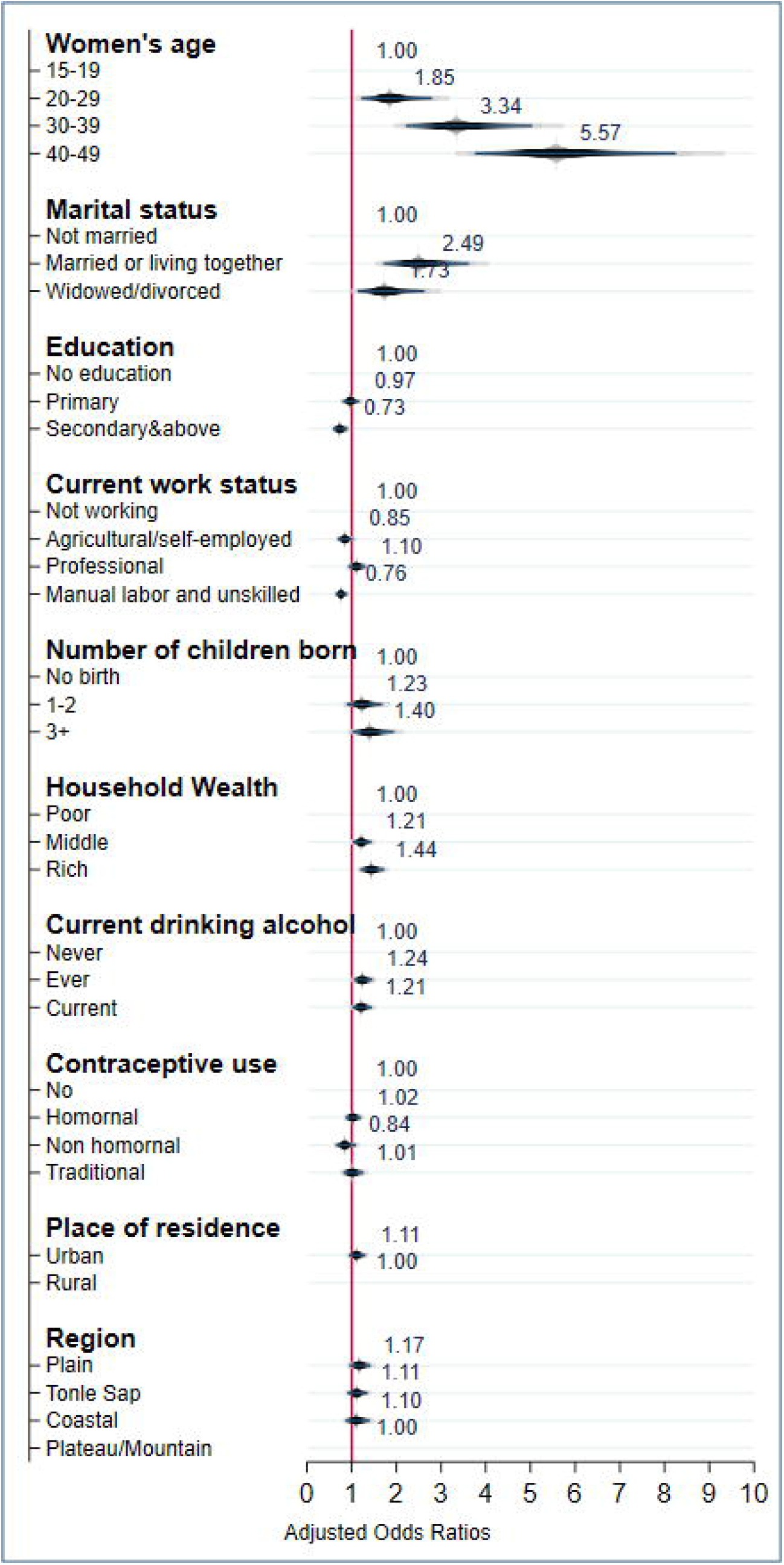
Adjusted analyses of the odds of women of reproductive age being overweight and obese in Cambodia **(n= 9**,**225)**.

## Discussion

The overall prevalence of overweight and obesity among WRA in Cambodia was 22.56% (95% CI: 21.5-23.7), and 5.61% (95% CI: 5.0-6.3), which has increased from the past survey estimates of overall overweight and obesity among women aged 15-49 years, from 6% in 2000 to 18% in 2014 [3], becoming a public health problem. Also, slightly consistent with the Global Nutrition Report 2019 showed that 26.1% of women were overweight, and obesity was 6.3% of women [6]. At the same time overweight and obesity is highest proportion among WRA resided in active economic provinces such as Kampong Cham, Kandal, Svay Rieng, Preah Sihanouk, Phnom Penh (**see Fig 1**). The variation and increase in overweight and obesity in Cambodia maybe explain by changes in lifestyles living condition, urbanization [11]. For example, the consumption of fast foods high in sugar, fat and salt has become widespread [11]. Further, physical activity has decreased due to improved transportation [11]. According to CDHS 2021-22 reported high prevalence of women consumed sweet beverages, and consumed unhealthy foods the previous day and drinking alcohol in past 30 days [8]. Factors associated with being overweight or obese for non-pregnant women include older age, increased parity, marriage and ever-married, middle, and rich household wealth index, and alcohol consumption. Women with higher education and those working in manual labor were less likely to be overweight or obese.

Older aged women were more likely to be overweight or obese as compared to younger women aged 15-19 years. This is consistent with other studies that showed obesity was more prevalent in older WRA [3,27]. The risk of women becoming overweight or obese rises with age, possibly due to unhealthy food consumption and a lack of physical activity [28]. Married women were more likely to be overweight or obese as compared to single women. Similar to previous study on the prevalence of overweight and obesity among WRA and its associated factors utilized CDHS data since 2014 [3], and in line with pooled analysis of DHS data among WRA in Bangladesh, Nepal, and India [29]. The explanation for that could be that married women busy working and take care their children and family may they do not pay more attention to nutritional and physical activity [3]. Women with better socioeconomic status and who live in urban areas had higher odds of being overweight and obese, which is consistent with prior research [30,31]. Women with higher socioeconomic status tend to use more improved technologies for a more comfortable lifestyle [19,32]. Women have at least three and more children at risk of being overweight and obese, similar to other studies [3,33]. During pregnancy, factors such as stress, depression, and anxiety may play a role in hypothalamic-pituitary-adrenal hyperactivity [34,35]. Women with several children may also have gained weight due to their reduced physical activity and have less time to focus on health behaviors, including weight management. In addition, women ever and currently consuming alcohol were significantly associated with higher risks of being overweight and obese. The association between alcohol consumption status and overweight and obesity has been studied thoroughly in various populations, such as Urban Cambodia and Hawassa City, southern Ethiopia [20,21]. Commonly, alcohol consumption is considered to increase appetite, resulting in excessive energy intake and leading to overweight and obesity [36].

In contrast, women with higher education are less likely to be overweight or obese than women with no school due to increased knowledge and awareness; for example, studies in Cambodia, Nigeria, and South Korea reported that educated women had a lower risk of being overweight or obese and that this may be linked to education influencing healthy behavior [3,37-40]. Education is a critical predictor of women’s healthy behaviors and health outcomes, including diet and physical activity [41]. Although overweight and obesity were not significantly associated with geographical regions of residence, those from the Plain and Coastal region had a high rate of overweight and obesity, 29.91% and 28.74%, respectively. Also, women who lived in urban areas had a higher prevalence of overweight and obesity, 29.90%. Prior research on overweight and obesity among women of reproductive age in Cambodia found women residing in urban had a significant association with overweight and obesity [3]; our findings suggest a need for more regionally representative studies.

### limitations and strengths

Our study has several limitations. First, because CDHS 2021-2022 collects cross-sectional data, our analysis could not explore changes over time. Second, data on several critical variables, such as women’s food-consumption behaviors, were not collected by CDHS, and CDHS did not objectively measure physical activity; therefore, our ability to examine the association of these variables with overweight and obesity was restricted. Third, numerous psychological factors (e.g., depressive and anxiety disorders) and physiological factors may also be associated with obesity. Still, these factors were not included in this study because they were unavailable in the data set. Finally, because the data were available only for women aged 15 to 49, our results may need to be generalizable to girls younger than 15 and women older than 49. Despite these limitations, our results contribute to the literature on the prevalence and association between socio-demographic and behavioral variables and the status of overweight and obesity among women of reproductive age in Cambodia. The major strength of this study is that it is the first to use nationally representative data with a high response rate of 97% to examine the prevalence and predictors of overweight and obesity in this country. Data were collected using validated survey methods, including calibrated measurement tools and highly trained data collectors, contributing to improved data quality [42]. Formally incorporating the complex survey design and sampling weights into the analysis bolsters the rigor of the analysis and enables generalizing our findings to the population of non-pregnant women in Cambodia.

## Conclusion

The overall prevalence of overweight and obesity among non-pregnant in Cambodia was very high. Women being older age, married, rich wealth indices, and alcohol consumption as risk factors associated with overweight and obesity. However, women being higher education levels, and manual labor and unskilled employment as reduce the risk of overweight and obesity among women of reproductive age in Cambodia. It is crucial to design intervention programs that target these socio-demographic factors for older, married, wealthier, high parity and drinker alcohol women who are more prone to be overweight and obesity through raise awareness of the importance of consuming healthy food and the benefits of regular physical activity, especially among older women monitor their weight, blood pressure and blood sugar with healthcare profession.

## Abbreviations

BMI,: Body mass index;
NCDs,: Non-communicable diseases;
WHO,: World Health Organization;
CDHS,: Cambodia Demographic Health Survey;
WRA,: Women reproductive age;
AOR,: Adjusted odds ratio;
PPS,: Probability proportional to size.
EA,: Enumeration areas

## Data Availability

Our study used the 2021-2022 Cambodia Demographic and Health Survey (CDHS) datasets. The DHS data are publicly available from the website at (URL:https://www.dhsprogram.com/data/available-datasets.cfm).

The Shapefiles for administrative boundaries in Cambodia are publicly accessible through the DHS website at (URL:https://spatialdata.dhsprogram.com/boundaries/#view=table&countryId=KH).

## Authors’ Contributions

**Samnang Um**. Contributed to conceptualization, and methods, performed the data analysis, interpreted the data, wrote the original draft, and reviewed/edited the manuscript. **Yom An**. Contributed to technical methods evaluation, support data analysis, and reviewing/editing the manuscript. Finally, all authors read and approved the final manuscript.

## Acknowledgments

The authors would like to thank DHS-ICF, who approved the data used for this paper. We want to acknowledge the management team of the National Institute of Public Health, Phnom Penh, Cambodia, namely Professor. **Chhea Chhorvann**. Professor. **Heng Sopheab** for their continued support and encouragement.

## Supporting information

**Table S1**. Results of checking multicollinearity using Variance Inflation Factor (VIF)

**Table S2**. Prevalence of overweight and obesity among women reproductive age, CDHS 2021-2022 (**n=9,417**)

## References

1. World Health Organization, Fact sheets, Obesity and overweight, Available from [https://www.who.int/news-room/fact-sheets/detail/obesity-and-overweightxs], Accessed on April 10, 2023.

2. Trends in adult body-mass index in 200 countries from 1975 to 2014: a pooled analysis of 1698 population-based measurement studies with 19·2 million participants. Lancet (London, England). 2016;387(10026):1377–96. Epub 2016/04/27. doi: 10.1016/s0140-6736(16)30054-x. PubMed PMID: 27115820.

3. Um S, Yom A J AM, Sopheab H. Overweight and obesity among women at reproductive age in Cambodia: Data analysis of Cambodia Demographic and Health Survey 2014. PLOS global public health. 2023;3(3):e0001768. Epub 2023/04/01. doi: 10.1371/journal.pgph.0001768. PubMed PMID: 37000710; PubMed Central PMCID: PMCPmc10065241.

4. Yu Z, Han S, Zhu J, Sun X, Ji C, Guo X. Pre-pregnancy body mass index in relation to infant birth weight and offspring overweight/obesity: a systematic review and meta-analysis. PLoS One. 2013;8(4):e61627. Epub 2013/04/25. doi: 10.1371/journal.pone.0061627. PubMed PMID: 23613888; PubMed Central PMCID: PMCPmc3628788.

5. Stanaway JD, Afshin A, Gakidou E, Lim SS, Abate D, Abate KH, et al. Global, regional, and national comparative risk assessment of 84 behavioural, environmental and occupational, and metabolic risks or clusters of risks for 195 countries and territories, 1990–2017: a systematic analysis for the Global Burden of Disease Study 2017. The Lancet. 2018;392(10159):1923–94.

6. Report GN. Country Nutrition Profiles, Available from; [https://globalnutritionreport.org/resources/nutrition-profiles/asia/south-eastern-asia/cambodia/]; Accessed on April 10, 2023.

7. National Institute of Statistics (NIS). (2020). General population census of the Kingdom of Cambodia 2019; accessed on November 4th 2021, availbale from: [https://www.nis.gov.kh/nis/Census2019/Final%20General%20Population%20Census%202019-English.pdf].

8. National Institute of Statistics (NIS) [Cambodia], Ministry of Health (MoH) [Cambodia], and ICF. 2023. Cambodia Demographic and Health Survey 2021–22 Final Report. Phnom Penh, Cambodia, and Rockville, Maryland, USA: NIS, MoH, and ICF.

9. World Food Programme in Cambodia; [https://www.wfp.org/countries/cambodia]: Accessed on April 10, 2023.

10. Balarajan Y, Villamor E. Nationally representative surveys show recent increases in the prevalence of overweight and obesity among women of reproductive age in Bangladesh, Nepal, and India. The journal of nutrition. 2009;139(11):2139–44.

11. Abrha S, Shiferaw S, Ahmed KY. Overweight and obesity and its socio-demographic correlates among urban Ethiopian women: evidence from the 2011 EDHS. BMC Public Health. 2016;16:1–7.

12. Yeshaw Y, Kebede SA, Liyew AM, Tesema GA, Agegnehu CD, Teshale AB, et al. Determinants of overweight/obesity among reproductive age group women in Ethiopia: multilevel analysis of Ethiopian demographic and health survey. BMJ open. 2020;10(3):e034963.

13. Oddo VM, Bleich SN, Pollack KM, Surkan PJ, Mueller NT, Jones-Smith JC. The weight of work: the association between maternal employment and overweight in low-and middle-income countries. International Journal of Behavioral Nutrition and Physical Activity. 2017;14(1):1–10.

14. Lopez LM, Bernholc A, Chen M, Grey TW, Otterness C, Westhoff C, et al. Hormonal contraceptives for contraception in overweight or obese women. Cochrane Database of Systematic Reviews. 2016;(8).

15. Aryeetey R, Lartey A, Marquis GS, Nti H, Colecraft E, Brown P. Prevalence and predictors of overweight and obesity among school-aged children in urban Ghana. BMC obesity. 2017;4:1–8.

16. de Munter Js, Tynelius P, Magnusson C, Rasmussen F. Longitudinal analysis of lifestyle habits in relation to body mass index, onset of overweight and obesity: results from a large population-based cohort in Sweden. Scandinavian journal of public health. 2015;43(3):236–45.

17. Piirtola M, Jelenkovic A, Latvala A, Sund R, Honda C, Inui F, et al. Association of current and former smoking with body mass index: A study of smoking discordant twin pairs from 21 twin cohorts. PloS one. 2018;13(7):e0200140.

18. Chiolero A, Jacot□Sadowski I, Faeh D, Paccaud F, Cornuz J. Association of cigarettes smoked daily with obesity in a general adult population. Obesity. 2007;15(5):1311–8.

19. Ghose B. Frequency of TV viewing and prevalence of overweight and obesity among adult women in Bangladesh: a cross-sectional study. BMJ open. 2017;7(1):e014399.

20. Dagne S, Gelaw YA, Abebe Z, Wassie MM. Factors associated with overweight and obesity among adults in northeast Ethiopia: a cross-sectional study. Diabetes, metabolic syndrome and obesity: targets and therapy. 2019:391–9.

21. Tamaoki M, Honda I, Nakanishi K, Nakajima M, Cheam S, Okawada M, et al. Lifestyle Factors Associated with Metabolic Syndrome in Urban Cambodia. International Journal of Environmental Research and Public Health. 2022;19(17):10481.

22. Darebo T, Mesfin A, Gebremedhin S. Prevalence and factors associated with overweight and obesity among adults in Hawassa city, southern Ethiopia: a community based cross-sectional study. BMC obesity. 2019;6:1–10.

23. World Health Organization, Fact sheets, A healthy lifestyle - WHO recommendations. Available from [https://www.who.int/europe/news-room/fact-sheets/item/a-healthy-lifestyle---who-recommendations], Accessed on April 10, 2023.

24. StataCorp. yv2021. Stata Statistical Software: Release 17. College Station, TX: StataCorp LLC. Availbale from: [https://www.stata.com/support/faqs/resources/citing-software-documentation-faqs/].

25. Bandoli G, Palmsten K, Chambers CD, Jelliffe-Pawlowski LL, Baer RJ, Thompson CA. Revisiting the Table 2 fallacy: A motivating example examining preeclampsia and preterm birth. Paediatr Perinat Epidemiol. 2018;32(4):390–7. Epub 20180521. doi: 10.1111/ppe.12474. PubMed PMID: 29782045; PubMed Central PMCID: PMCPMC6103824.

26. Westreich D, Greenland S. The table 2 fallacy: presenting and interpreting confounder and modifier coefficients. Am J Epidemiol. 2013;177(4):292–8. Epub 20130130. doi: 10.1093/aje/kws412. PubMed PMID: 23371353; PubMed Central PMCID: PMCPMC3626058.

27. Biswas T, Uddin MJ, Mamun AA, Pervin S P Garnett S. Increasing prevalence of overweight and obesity in Bangladeshi women of reproductive age: Findings from 2004 to 2014. PloS one. 2017;12(7):e0181080.

28. Alemu E, Atnafu A, Yitayal M, Yimam K. Prevalence of overweight and/or obesity and associated factors among high school adolescents in Arada Sub city, Addis Ababa, Ethiopia. Journal of nutrition & food sciences. 2014;4(2):1.

29. Balarajan Y, Villamor E. Nationally representative surveys show recent increases in the prevalence of overweight and obesity among women of reproductive age in Bangladesh, Nepal, and India. J Nutr. 2009;139(11):2139–44. Epub 20090923. doi: 10.3945/jn.109.112029. PubMed PMID: 19776182.

30. Mangemba NT, San Sebastian M. Societal risk factors for overweight and obesity in women in Zimbabwe: a cross-sectional study. BMC public health. 2020;20(1):1–8.

31. Kamal SM, Hassan CH, Alam GM. Dual burden of underweight and overweight among women in Bangladesh: patterns, prevalence, and sociodemographic correlates. Journal of health, population, and nutrition. 2015;33(1):92.

32. Islam M, Sathi NJ, Abdullah HM, Naime J, Butt ZA. Factors Affecting the Utilization of Antenatal Care Services During Pregnancy in Bangladesh and 28 Other Low-and Middle-income Countries: A Meta-analysis of Demographic and Health Survey Data. Dr Sulaiman Al Habib Medical Journal. 2022;4(1):19–31.

33. Mbochi RW, Kuria E, Kimiywe J, Ochola S, Steyn NP. Predictors of overweight and obesity in adult women in Nairobi Province, Kenya. BMC public health. 2012;12(1):1–9.

34. Li W, Wang Y, Shen L, Song L, Li H, Liu B, et al. Association between parity and obesity patterns in a middle-aged and older Chinese population: a cross-sectional analysis in the Tongji-Dongfeng cohort study. Nutrition & metabolism. 2016;13(1):1–8.

35. Pasquali R, Vicennati V, Cacciari M, Pagotto U. The hypothalamic□pituitary□adrenal axis activity in obesity and the metabolic syndrome. Annals of the New York Academy of Sciences. 2006;1083(1):111–28.

36. Yeomans MR. Alcohol, appetite and energy balance: is alcohol intake a risk factor for obesity? Physiology & behavior. 2010;100(1):82–9. Epub 2010/01/26. doi: 10.1016/j.physbeh.2010.01.012. PubMed PMID: 20096714.

37. Hamad R, Elser H, Tran DC, Rehkopf DH, Goodman SN. How and why studies disagree about the effects of education on health: A systematic review and meta-analysis of studies of compulsory schooling laws. Social Science & Medicine. 2018;212:168–78.

38. Chung A, Backholer K, Wong E, Palermo C, Keating C, Peeters A. Trends in child and adolescent obesity prevalence in economically advanced countries according to socioeconomic position: a systematic review. Obesity reviews. 2016;17(3):276–95.

39. Barlow P. The effect of schooling on women’s overweight and obesity: A natural experiment in Nigeria. Demography. 2021;58(2):685–710.

40. McLaren L. Socioeconomic status and obesity. Epidemiologic reviews. 2007;29(1):29–48.

41. Liu H, Guo G. Lifetime socioeconomic status, historical context, and genetic inheritance in shaping body mass in middle and late adulthood. American sociological review. 2015;80(4):705–37.

42. Bowler P. CUNNINGHAM, A. and Jardine, N.(editors). Romanticism and the sciences. Cambridge University Press, Cambridge: 1990. Pp xxii, 345; illustrated. Price:£ 15, US $19.95. ISBN: 0-521-35685-7 (paperback). 1992.

