## Supplementary material for "Factors Associated with Overweight and Obesity among Women of Reproductive Age in Cambodia: Analysis of Cambodia Demographic and Health Survey": S1 Table

**Supporting information**

**S1 Table**. Results of checking multicollinearity using Variance Inflation Factor (VIF)

| **Variables** | **VIF** |
| --- | --- |
| Woman’s age | 1.79 |
| Number of children born | 1.68 |
| Marital status | 1.54 |
| Household wealth index | 1.38 |
| Place of residence | 1.28 |
| Contraceptive use | 1.19 |
| Education | 1.15 |
| Region | 1.05 |
| Alcohol consumption | 1.03 |
