## Supplementary material for "Factors Associated with Overweight and Obesity among Women of Reproductive Age in Cambodia: Analysis of Cambodia Demographic and Health Survey": S2 Table

**Supporting information**

**S2 Table**. Prevalence of overweight and/or obesity among women of reproductive age, CDHS 2021-2022 **(n=9,417)**

| **Provinces** | **Number of women** | **% Overweight and/or obese** | **95% CI** | **% Normal and/or Underweight** | **95% CI** |
| --- | --- | --- | --- | --- | --- |
| Banteay Meanchey | 365 | 25.2 | [20.6-30.5] | 74.8 | [69.5-79.4] |
| Battambang | 650 | 28.1 | [24.4-32.0] | 71.9 | [68.0-75.6] |
| Kampong Cham | 560 | **34.1** | [29.2-39.4] | 65.9 | [60.6-70.8] |
| Kampong Chhnang | 330 | 14.6 | [11.9-17.7] | 85.4 | [82.3-88.1] |
| Kampong Speu | 601 | 26.8 | [22.1-32.0] | 73.2 | [68.0-77.9] |
| Kampong Thom | 395 | 30.7 | [25.7-36.2] | 69.3 | [63.8-74.3] |
| Kampot | 383 | 28.6 | [24.0-33.7] | 71.4 | [66.3-76.0] |
| Kandal | 680 | **32.3** | [27.2-38.0] | 67.7 | [62.0-72.8] |
| Koh Kong | 68 | 26.0 | [20.6-32.1] | 74.0 | [67.9-79.4] |
| Kratie | 213 | 27.6 | [23.1-32.5] | 72.4 | [67.5-76.9] |
| Mondul Kiri | 56 | 25.3 | [20.5-30.7] | 74.7 | [69.3-79.5] |
| Phnom Penh | 1,573 | 30.3 | [25.8-35.3] | 69.7 | [64.7-74.2] |
| Preah Vihear | 154 | 24.9 | [18.9-32.0] | 75.1 | [68.0-81.1] |
| Prey Veng | 573 | 26.2 | [21.4-31.6] | 73.8 | [68.4-78.6] |
| Pursat | 220 | 30.4 | [25.0-36.4] | 69.6 | [63.6-75.0] |
| Ratanak Kiri | 137 | **9.4** | [6.8-12.8] | 90.6 | [87.2-93.2] |
| Siemreap | 742 | 29.2 | [24.0-35.1] | 70.8 | [64.9-76.0] |
| Preah Sihanouk | 118 | **31.2** | [27.1-35.5] | 68.8 | [64.5-72.9] |
| Stung Treng | 93 | 22.1 | [18.0-26.8] | 77.9 | [73.2-82.0] |
| Svay Rieng | 357 | **32.0** | [27.4-37.1] | 68.0 | [62.9-72.6] |
| Takeo | 555 | 23.8 | [20.0-27.9] | 76.2 | [72.1-80.0] |
| Oddar Meanchey | 114 | 23.6 | [19.7-27.9] | 76.4 | [72.1-80.3] |
| Kep | 27 | 25.3 | [20.9-30.2] | 74.7 | [69.8-79.1] |
| Pailin | 45 | 30.7 | [26.6-35.2] | 69.3 | [64.8-73.4] |
| Tboung Khmum | 411 | 30.3 | [25.8-35.1] | 69.7 | [64.9-74.2] |
